# A National Survey Exploring Dementia Diagnostics and Care Provisions in Primary Care in England

**DOI:** 10.64898/2026.09.11.26362825

**Authors:** Ayoni Darby, Jasmine Shaw, Claudia Cooper, Natalia Chemas, Charles R Marshall, Naaheed Mukadam, Kate Smith, Rozeena Khan, Sube Banerjee, Greta Rait

**Affiliations:** Queen Mary University of London; University College London; Queen Mary University of London PPI; University of Nottingham

**Keywords:** Dementia, general practice, primary care, disease modifying treatments, England WC:3, GGG

## Abstract

**Background:** The 10 Year Health Plan for England indicates governmental intention to shift care from hospital to community. Disease Modifying Treatments (DMTs) and biomarker tests for dementia will likely be introduced, with potentially significant impacts on community care, particularly general practice.

**Aim:** To explore dementia diagnosis, post diagnostic support and expectations around future care in primary care settings.

**Design and Setting:** Codesigned survey with general practitioners (GPs), policy researchers, carers and people with lived experience. One GP per practice invited from five of England’s Integrated Care Boards (ICBs).

**Methods:** Inductive content analysis for qualitative responses and non-weighted means calculated for quantitative answers. Findings contextualised using national data.

**Results:** Between September and December 2025, 80/1094 (7.3%) eligible practices participated (80 GPs). Most, (79/80, 98.8%) used cognitive assessments and 100% (80/80) blood tests to support dementia diagnosis, with 27/70 (39.7%) able to order brain scans directly. GPs primarily conducted annual physical, mental or care plan reviews for people with known dementia. Respondents considered it unlikely (47/80, 58.8%) they would formally diagnose dementia or assess (50/80, 62.3%) for DMT eligibility in the next five years. GPs considered it somewhat/most likely (54/80, 67.5%) they could offer blood biomarker tests and continue established DMT prescribing or monitoring (61/80, 76.3%). Resource, capacity and training needs were perceived barriers.

**Conclusion:** GPs are the first point of contact for accessing a dementia diagnosis in England. Accommodating novel diagnostic tools and treatments requires a reimagining of the relationship between primary, secondary and social care to address GP reported constraints.

## Introduction

Dementia prevalence is rising worldwide; it is one of the most pressing global healthcare concerns. The number of people with a formal diagnosis of dementia in England increased from 427,176 in 2022 to 498,926 in 2025 and it is the leading cause of death in England (1,2). Eliciting profound personal impact on the individual and their families, dementia also causes wider effects on the National Health Service (NHS), social care and society, with UK annual costs expected to reach £90 billion by 2040 (3).

“The 10 Year Health Plan for England” (4) asks health and social care services to deliver three “big shifts” over the next decade: from sickness to prevention, hospital to community and analogue to digital. Primary care will be central to all of these, and though necessary, they will be delivered in challenging times. The number of fully qualified General Practitioners (GPs) is falling, with 773 fewer full-time GPs in 2025 compared to 2015 (5). This is whilst the average number of appointments provided has increased with a record 380 million delivered between June 2024 to 2025 (6). Nearly two thirds (63%) of GPs feel they have insufficient appointment times to build patient relationships and deliver quality care (5).

These constraints have implications for people affected by dementia, particularly as services are already stretched. A fifth of UK people wait over two years for a dementia diagnosis following their first GP consultation (7). This matters because early identification improves quality of life and diminishes health and care expenditure by reducing hospitalisations and care home admissions (8,9). In the UK, GPs are customarily the first practitioners consulted about cognitive changes, so they have a critical role in enabling early diagnosis by promptly referring those likely to have dementia to secondary care and diverting those unlikely to have the disorder to other, more appropriate pathways (10).

GPs are typically involved in dementia post-diagnostic care (11). Memory clinics introduced nationwide in 2009 have helped alleviate pressure on primary care by diagnosing and providing symptomatic treatments to individuals with dementia, with the proportion of people diagnosed doubling between 2005 and 2015 (12,13). However, after initial post-diagnostic encounters, patients including those with complex existing or emerging comorbidities and with neuropsychiatric or social health concerns typically have their treatment devolved back to primary care with the support of secondary or community mental health services.

Recent primary care research evaluating GP attitudes towards existing Alzheimer’s pathways in England discovered a self-reported skills gap (14). Practitioners vocalised confidence and capacity concerns around prescribing the already established symptomatic Alzheimer’s drugs, acetylcholinesterase inhibitors and memantine.

The emergence of novel pharmacological interventions means the first efficacious disease modifying treatments (DMTs) could soon be approved for NHS use (15).

Pressures on healthcare services may increase with primary care clinicians required to facilitate prompt and accurate dementia diagnoses, potentially through novel biomarkers. As the previous licensing of DMTs for other conditions such as multiple sclerosis (MS) was followed by reports of disparities in eligible patients’ ability to access these medications planning is needed for successful dementia DMT integration (16,17).

A 2023 study amongst a large German cohort surveyed 4,511 GPs, identifying time and funding to be the primary impediments to timely diagnosis (18). In the most recent English survey dated 2010, half of GP respondents expressed a lack of satisfactory specialist services and highlighted a dearth of sufficient dementia training (19). This current study sought to be the first in 15 years to recruit a national sample of GPs to ascertain contemporary diagnostic frameworks, provisions and views on system preparedness within primary care in the context of advancing therapies, an increased awareness of dementia and the inception of several national healthcare strategies.

## Objectives

Embedded within a broader national research project exploring the overall landscape of dementia provisions within differing health and social care organisations (20,21), we surveyed GPs in diverse areas of England. Our objectives were to:

1. Explore the procedures of primary care provision pre- and post-diagnosis of dementia, including practice access to diagnostic tools, allocation of diagnostic support, and reliance on community and secondary care support.
2. Gather GP expectations of future primary care services for dementia, including perceived barriers and facilitators for diagnostic responsibilities, DMT related responsibilities, and desired staff training.
3. Contextualise findings by comparing the characteristics of participating Integrated Care Boards (ICBs) to national public health care data.

## Methodology

This cross-sectional national provider survey was conducted by the NIHR Dementia and Neurodegeneration Policy Research Unit at Queen Mary, University of London (DeNPRU-QM), commissioned by the Department of Health and Social Care (DHSC) and approved by National Research Ethics service and Health Research Authority on 14.05.2024 (24/IEC08/0008).

### Survey development and co-design

We developed the survey iteratively using a co-design approach with members of the public with lived experience of dementia, and GPs working in diverse regions of England, all of whom received a monetary voucher in recognition of their contributions to this research.

The Patient and Public Involvement and Experience (PPIE) group at DeNPRU-QM met with researchers in an online focus group, contributing to survey development. This was facilitated by two members of the DeNPRU-QM team (CK and JS) with seven PPIE members: four with direct experience of dementia (living with a diagnosis or a family carer) and three with a diagnosis of MS and experience accessing DMTs. The survey was drafted reflecting the aims of the study and input from PPIE members.

In three online meetings, two researchers (GR and JS) met with: 1) one PCN (Primary Care Network) Clinical Director and GP 2) one GP, and 3) three GPs. Before these meetings, participants were sent an evidence pack to provide context and the drafted survey. During the meetings, members proposed and discussed survey items in line with the evidence and aims, providing feedback using a structure, process and outcome framework. After the GP co-design groups, the drafted survey was reviewed reflecting participants comments before the final version was produced using Qualtrics (www.qualtrics.com).

The focal points of the survey were what pre and post diagnostic support was offered by practices and perceived readiness for the introduction of DMTs. In the final iteration questions asked about specific diagnostic tools and assessments used by services, alternate post diagnostic support providers, self-reported competence at diagnosing, provisions for other cognitive dysfunction including Mild Cognitive Impairment (MCI), functional memory complaints and cognitive impairment in people with Parkinson’s disease, expectations for the future of dementia care, participant demographics and service characteristics. The survey consisted of descriptive, discrete choice closed questions and open questions with free text responses for further expansion (survey in supplementary material).

### Survey sample and data collection

We selected five geographically diverse ICBs in England within which to conduct the survey, in London, Midlands, South East, and two in North East and Yorkshire. We selected ICBs for ethnic and deprivation diversity and geographical heterogeneity with urban, rural and coastal representation. We used publicly available national health data via the government Fingertips tool to identify the 1,094 eligible GP practices (22).

Participants were GPs currently employed by practices within the five specified ICBs. Our intention *a priori* was to purposively select GPs from 75 of these practices to reflect diversity of age, gender and practice size. In practice, due to the low response rate, we contacted most eligible surgeries. Contact was through the National Institute for Health and Care Research (NIHR) Research Delivery Networks (RDN) or direct by AD. Practices were sent a cover letter, participant information sheet (PIS) and link to complete the survey independently online via Qualtrics, or if they preferred on the phone with a researcher. Consent was completed electronically prior to survey completion. No renumeration was provided for completion. We only included one GP from each practice.

### Contextual national data

To contextualise our findings, we compared indicators of the chosen ICBs to national figures. The co-design group thought obtaining publicly available data would increase the response rate by reducing participating GP burden. We accessed Fingertips data on 15^th^ January 2026. Using the public health profiles, we extracted the number of registered persons per practice, GP performance data, dementia prevalence and deprivation indices at ICB and nationwide level. To obtain ethnicity estimates, we used the application programming interface then imported data to R Studio Team (23), checking for missing values and excluding null entries. Unweighted means were calculated across GP practices to derive ICB level estimates. For inactive indicators (total funding per patient and number of full-time workers) the DHSC provided the latest 2021 figures which were read on R, with null entries excluded and unweighted mean values procured.

### Analysis

We exported survey data from Qualtrics to Microsoft Excel (24). We calculated the mean age of participants and number of patients seen per annum on RStudio Team.

We used standard descriptive statistics to summarise responses to discrete choice items; where items described a range, the midpoint in analyses was used (e.g. 5-10 became 7.5) and if greater or less than signs were written a minimum estimate was assigned (for example >20 became 21).

We processed free text responses in Microsoft Excel, conducting inductive qualitative content analysis. AD coded all responses, developing a framework. CC then independently coded 10% of responses for each question, using the frame, discussing any discrepancies in codes and iteratively modifying the coding frame. We used frequency counts calculated to summarise the prevailing themes amongst those that had written narrative comments with some quotes fitting multiple codes. As open-ended responses were optional, the qualitative coding was not representative of all participants, but highlighted the salient points identified by a select sample that gave supplementary information. Only original narrative comments were coded, with "as/see above" comments omitted.

## Results

### Sample description (*Table 1*)

Between September and December 2025, 140 physicians accessed the survey link with 86 completed submissions (54 blank questionnaires were excluded), from which we included 80/86 responses. Three responses were removed as participants had omitted their ICB and three were from practices from which a response had already been received (only the first entry was accepted).

**Table 1:**
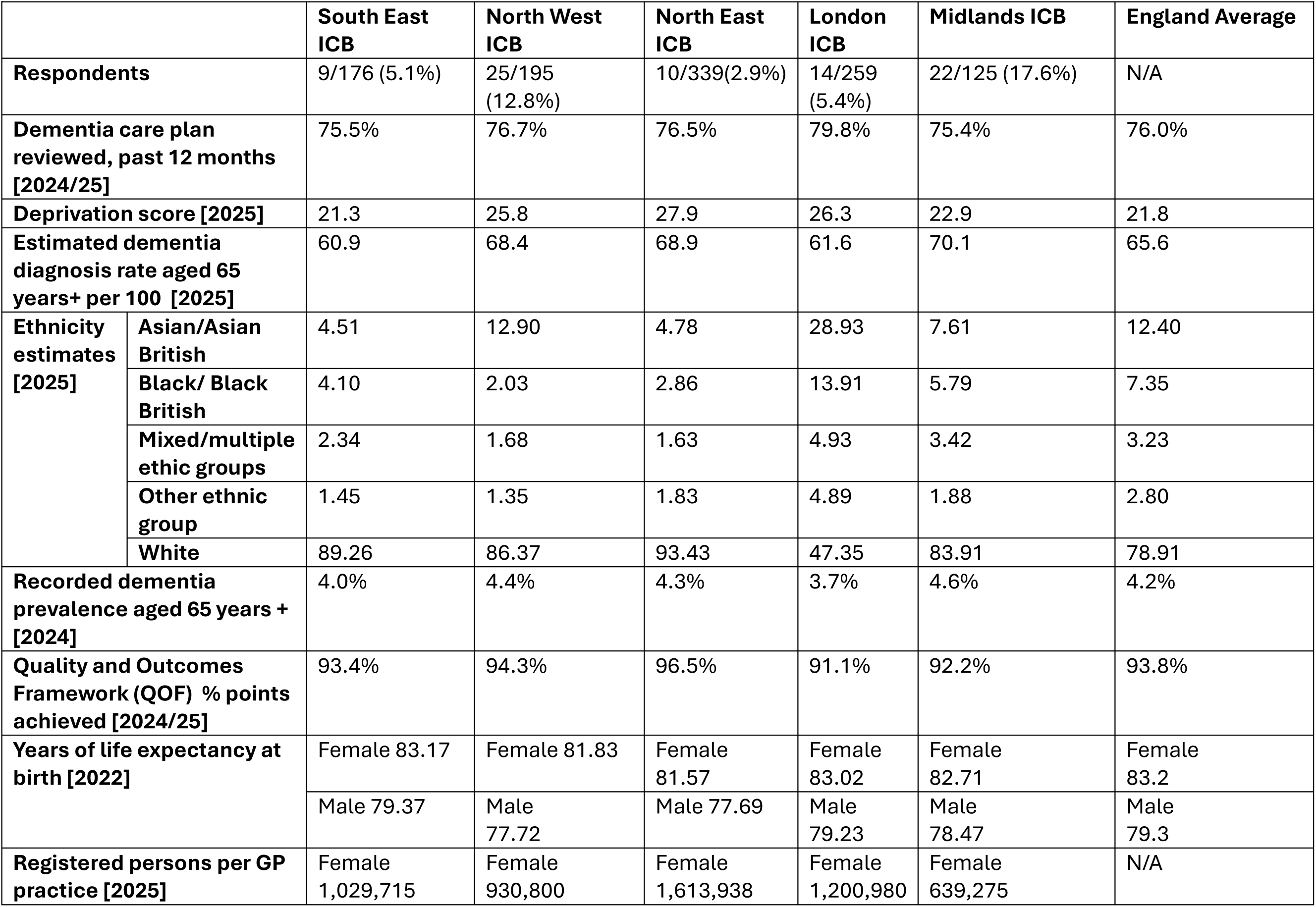
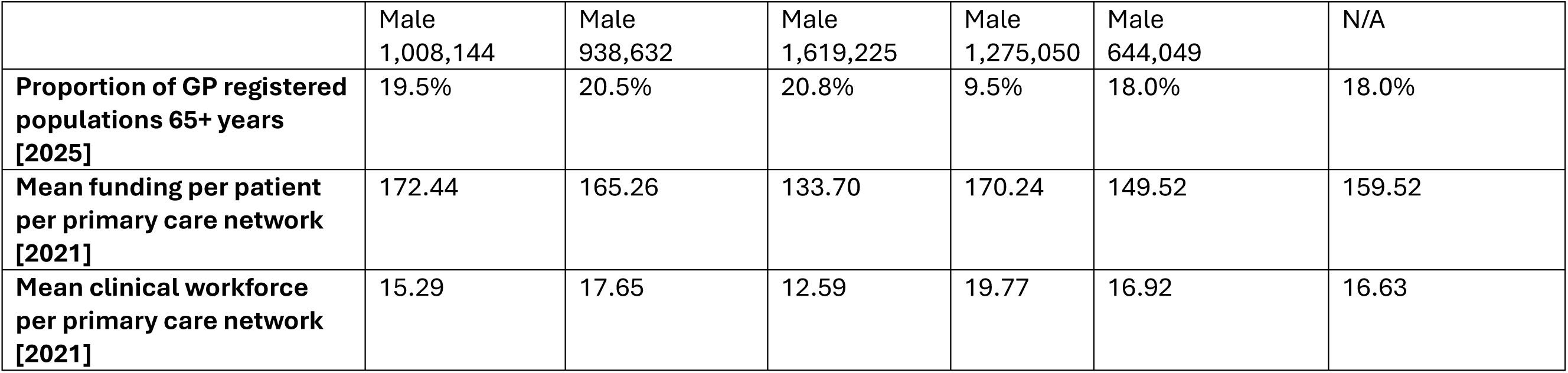
Proportion of practices that responded per ICB and descriptive statistics of each ICB.

|  |  |  |  |  |  |  |  |
| --- | --- | --- | --- | --- | --- | --- | --- |
|  |  | South East ICB | North West ICB | North East ICB | London ICB | Midlands ICB | England Average |
| Respondents |  | 9/176 (5.1%) | 25/195 (12.8%) | 10/339(2.9%) | 14/259 (5.4%) | 22/125 (17.6%) | N/A |
| Dementia care plan reviewed, past 12 months [2024/25] |  | 75.5% | 76.7% | 76.5% | 79.8% | 75.4% | 76.0% |
| Deprivation score [2025] |  | 21.3 | 25.8 | 27.9 | 26.3 | 22.9 | 21.8 |
| Estimated dementia diagnosis rate aged 65 years+ per 100 [2025] |  | 60.9 | 68.4 | 68.9 | 61.6 | 70.1 | 65.6 |
| Ethnicity estimates [2025] | Asian/Asian British | 4.51 | 12.90 | 4.78 | 28.93 | 7.61 | 12.40 |
|  | Black/ Black British | 4.10 | 2.03 | 2.86 | 13.91 | 5.79 | 7.35 |
|  | Mixed/multiple ethic groups | 2.34 | 1.68 | 1.63 | 4.93 | 3.42 | 3.23 |
|  | Other ethnic group | 1.45 | 1.35 | 1.83 | 4.89 | 1.88 | 2.80 |
|  | White | 89.26 | 86.37 | 93.43 | 47.35 | 83.91 | 78.91 |
| Recorded dementia prevalence aged 65 years + [2024] |  | 4.0% | 4.4% | 4.3% | 3.7% | 4.6% | 4.2% |
| Quality and Outcomes Framework (QOF) % points achieved [2024/25] |  | 93.4% | 94.3% | 96.5% | 91.1% | 92.2% | 93.8% |
| Years of life expectancy at birth [2022] |  | Female 83.17 | Female 81.83 | Female 81.57 | Female 83.02 | Female 82.71 | Female 83.2 |
|  |  | Male 79.37 | Male 77.72 | Male 77.69 | Male 79.23 | Male 78.47 | Male 79.3 |
| Registered persons per GP practice [2025] |  | Female 1,029,715 | Female 930,800 | Female 1,613,938 | Female 1,200,980 | Female 639,275 | N/A |

|  |  |  |  |  |  |  |
| --- | --- | --- | --- | --- | --- | --- |
|  | Male<br>1,008,144 | Male<br>938,632 | Male<br>1,619,225 | Male<br>1,275,050 | Male<br>644,049 | N/A |
| **Proportion of GP registered**<br>**populations 65+ years**<br>**[2025]** | 19.5% | 20.5% | 20.8% | 9.5% | 18.0% | 18.0% |
| **Mean funding per patient**<br>**per primary care network**<br>**[2021]** | 172.44 | 165.26 | 133.70 | 170.24 | 149.52 | 159.52 |
| **Mean clinical workforce**<br>**per primary care network**<br>**[2021]** | 15.29 | 17.65 | 12.59 | 19.77 | 16.92 | 16.63 |

Figure 1 (supplementary material) shows the geographical distribution of participating practices and Table 1 the response rates. The overall response rate was 80/1094 (7.3%). Table 1 shows that four ICBs had slightly higher deprivation scores than national average and one had a lower value. Three areas had higher estimated dementia diagnosis rates and two lower rates than the England average. One urban ICB was the most ethnically diverse, with the four others recording White ethnic groups as more prevalent than national average.

### Description of participating practices and respondents (*Table 2*)

Table 2 describes participants and their GP practices. Respondent ethnicity, age and gender profiles were broadly reflective of the UK GP working population (25). The average length of time respondents had worked as a GP was 14.5 years (range: 0-40 years).

**Table 2:**
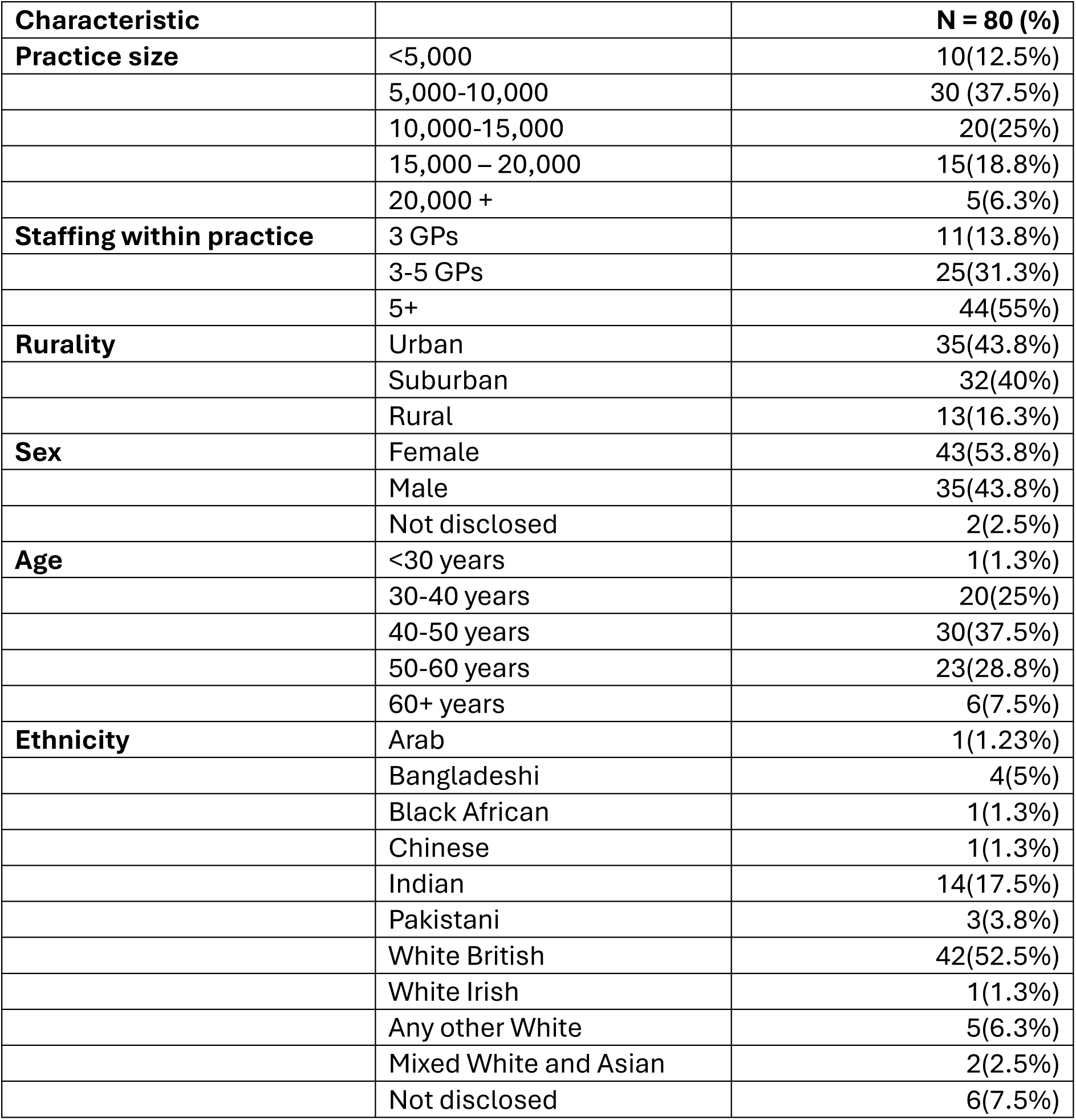
Personal demographics and practice characteristics of respondents.

|  |  |  |
| --- | --- | --- |
| **Characteristic** |  | **N = 80 (%)** |
| **Practice size** | <5,000 | 10(12.5%) |
|  | 5,000-10,000 | 30 (37.5%) |
|  | 10,000-15,000 | 20(25%) |
|  | 15,000 – 20,000 | 15(18.8%) |
|  | 20,000 + | 5(6.3%) |
| **Staffing within practice** | 3 GPs | 11(13.8%) |
|  | 3-5 GPs | 25(31.3%) |
|  | 5+ | 44(55%) |
| **Rurality** | Urban | 35(43.8%) |
|  | Suburban | 32(40%) |
|  | Rural | 13(16.3%) |
| **Sex** | Female | 43(53.8%) |
|  | Male | 35(43.8%) |
|  | Not disclosed | 2(2.5%) |
| **Age** | <30 years | 1(1.3%) |
|  | 30-40 years | 20(25%) |
|  | 40-50 years | 30(37.5%) |
|  | 50-60 years | 23(28.8%) |
|  | 60+ years | 6(7.5%) |
| **Ethnicity** | Arab | 1(1.23%) |
|  | Bangladeshi | 4(5%) |
|  | Black African | 1(1.3%) |
|  | Chinese | 1(1.3%) |
|  | Indian | 14(17.5%) |
|  | Pakistani | 3(3.8%) |
|  | White British | 42(52.5%) |
|  | White Irish | 1(1.3%) |
|  | Any other White | 5(6.3%) |
|  | Mixed White and Asian | 2(2.5%) |
|  | Not disclosed | 6(7.5%) |

### Diagnostic provisions (*Table 3*)

Responding GPs reported a non-weighted means average of consulting with 9.7 (range 0-78) patients annually, where the primary appointment reason were symptoms resulting in a suspected dementia diagnosis. Just less than a third used active monitoring involving listening, empathetic understanding and problem solving as part of their approach to dementia diagnosis. Almost all used cognitive tests to ascertain likely dementia cases, with the Six Item Cognitive Impairment (6-CIT) and General Practitioner Assessment of Cognition (GPCOG) most common. All respondents requested screening blood tests for metabolic causes of cognitive impairment as part of assessment, with two indicating that they had access to blood biomarkers. Under half of respondents were able to directly request imaging, twelve respondents did so regularly (7 requesting MRI, 5 CT head scans, but no PET scans). In terms of onward referrals, most (69/80, 86.3%) referred to memory services and 16/80 (20%) to older people’s community mental health teams, 6/80 (7.5%) neurology or 5/80 (6.3%) geriatrics.

**Table 3:** Diagnostic methods used at each practice.

| **Table 3: Diagnostic methods used at each practice** |  |  |
| --- | --- | --- |
| **Diagnostic method** |  | **Number of responses**<br>**(% that responded)** |
| **Active Monitoring (n=80)** |  | 23 (28.8%) |
| **Cognitive Screening/**<br>**Psychometric Tests used**<br>**(n=80)** | At least one test | 79 (98.8%) |
|  | 6-CIT | 32 (40.0%) |
|  | GPOG | 29 (36.3%) |
|  | ACE-III | 2 (2.5%) |
|  | MoCA | 5 (6.3%) |
|  | MMSE | 7 (8.8%) |
|  | Other | 2(2.5) |
| **Blood Tests (n=80)** |  | 80 (100%) |
| **Direct access to blood**<br>**biomarkers(n=80)** | No | 78(97.5%) |
|  | Yes | 2(2.5%) |
| **Direct access to**<br>**neuroimaging (n=68)** | Yes | 27(39.7%) |
|  | No | 41(60.3%) |

### Healthcare reviews (*Table 4*, Supplementary Table 1)

Most GPs conducted care plan (88.8%), mental health (83.8%), medication (81.3%) and physical health reviews (90%) for people with dementia annually, aligning with the latest guidance (26). Supplementary Table 1 shows content analysis results of free text responses. Many practices adopted a “*very patient specific*” approach responsive to individual patient needs (25/31, 80.6%) or the presence of other conditions (8/31, 25.8%) highlighting “*physical health reviews are only if they have other comorbidities like diabetes”* or that these were more frequent depending on the patient setting (6/31, 19.4%) such as a care home or hospital.

**Table 4:** For patients with dementia the frequency GP practices conduct reviews.

|  |  |  |  |  |
| --- | --- | --- | --- | --- |
|  | **Never**<br>**(n=80)** | **3 Monthly**<br>**(n=80)** | **6 Monthly**<br>**(n=80)** | **Yearly**<br>**(n=80)** |
| **Care Plan** | 4(5%) | 0(0%) | 5(6.3%) | 71(88.8%) |
| **Mental Health** | 6(7.5%) | 0(0%) | 7(8.8%) | 67(83.8%) |
| **Medication** | 1(1.3%) | 2(2.5%) | 12(15%) | 65(81.3%) |
| **Physical**<br>**Health** | 5(6.3%) | 0(0%) | 3(3.8%) | 72(90%) |
| **MDTs** | 52(65%) | 9(11.3%) | 3(3.8%) | 16(20%) |

Around a third (35%) of practices conducted multidisciplinary team meetings at least yearly. Some practices had regularly scheduled reviews (10.3%) including a “*monthly PCN MDT meeting run by a dementia nurse*” whilst others scheduled appointments in response to concerns.

### Post diagnostic care (Supplementary Table 2)

GPs collaborated with a range of dementia post diagnostic support providers: voluntary and community organisations (52/80, 65%), clinical memory services including psychiatry, neurology or geriatric specialists (50/80, 63%), social care services (46/80,57.5%), care homes (38/80,47.5%) and older adult mental health services (37/80,46.3%). A small number (7/80, 8.8%) stated that no other organisations were routinely engaged.

A minority of practices (10/80, 12.5%) offered specific services for people with dementia belonging to minority ethnic groups, including those with English as second language. These comprised social prescribers, interpreters, multilingual members of staff or support groups.

### Other cognitive presentations (Supplementary Table 3)

Over a quarter (21/80, 26.3%) of GPs surveyed worked within a practice that offered targeted signposting or support for people with MCI. This was in collaboration with third sector providers, including Age UK and Dementia UK, older peoples mental health teams, memory clinics, social prescribers and specialised carers support.

A greater proportion (31/80, 38.8%) of practices recorded specific signposting or tailored support in place for people with Parkinson’s disease who were experiencing cognitive impairment. These consisted of third sector workers or support groups (21.3%), Parkinson’s nurses (10%) and specialist teams including movement disorder clinics.

We asked what specific resources are available to support clinicians in managing subjective or functional memory complaints. National Institute for Health and Care Excellence (NICE)/Clinical Knowledge Summaries (CKS) were used by over a third of participants (30/71, 42.3%), assistance from specialist individuals or teams such as memory clinics and old age psychiatrists was available to 23.9% (17/71) alongside unspecified general training resources (15/71, 23.9%) whilst almost a third (20/71, 28.2%) were unaware of any specific resources or reported that very little was available, with no regional differences evident.

### Anticipated developments in the next five years

Supplementary Table 4 shows findings from our content analysis of free text responses, and Table 5 responses to survey questions regarding anticipated developments.

**Table 5:** In the next 5 years do GPs expect their practice to consider.

| **Table 5: In the next 5 years do GPs expect their practice to consider** |  |  |  |  |
| --- | --- | --- | --- | --- |
|  | **Not Likely** | **Somewhat**<br>**Likely** | **Most Likely** | **We Already**<br>**Do This** |
| **Formally diagnosing**<br>**dementia (n=80)** | 47(58.8%) | 23(28.8%) | 9(11.3%) | 1(1.3%) |
| **Taking bloods for**<br>**biomarker testing(n=80)** | 24(29.5%) | 37(46.3%) | 17(21.3%) | 2(2.5%) |
| **Assessing patient**<br>**eligibility for DMTs for**<br>**dementia (n=80)** | 50(62.3%) | 23(28.8%) | 7(8.8%) | Not<br>applicable |
| **Prescribing DMTs for**<br>**dementia (n=80)** | 26(32.5%) | 39(48.8%) | 15(18.8%) | Not<br>applicable |
| **Monitoring progress &**<br>**side effects of DMTs for**<br>**dementia (n=80)** | 19(23.8%) | 41(51.3%) | 20(25%) | Not<br>applicable |

#### Diagnosing dementia

Most GPs considered it unlikely that their practices would formally contemplate diagnosing dementia in the next five years (Table 5). Supplementary Table 4 shows that as for all innovations discussed, lack of confidence, time and capacity were the prevailing reasons cited.

The most frequently endorsed code (32/70, 45.7%), reflected in the response of those who felt their practice would not start to diagnose dementia, was “*lack of confidence, expertise and training*” with an understanding that dementia is “*a complex diagnosis that can be mimicked by other mental and physical health problems. It is a diagnosis made after a number of appointments and use of longer memory assessments than we are able to do in primary care*”. Other frequent codes explaining why diagnosis rates in primary care would not increase were a “lack of capacity and funding” (21/70, 30%), and a lack of diagnostic facilities (9/70, 12.9%). Policy around a shift to primary care was cited by 15/70 (22.9%) of respondents that considered it likely diagnosis in primary care would increase.

#### Introducing biomarkers (Supplementary Table 5)

Almost half of GPs (37/80, 43.3%) felt their practices would be somewhat likely to take bloods for specific dementia biomarker testing. One respondent commented that it would be a “*relatively easy change to make to our practice*” and that it would be “*very much welcomed as it will give us solid evidence to diagnose and treat within primary care*” echoed by 25/57 of free text responses (43.9%) that either expressed a willingness to utilise or felt biomarkers could become a part of routine screening. With requisite funding and access (20/57, 35.1%) and support or shared care (11/57. 19.3%) GPs posited blood biomarkers would likely be introduced. Conversely funding and capacity (8/57, 14.0%) issues were identified as reasons implementation would be unlikely with GP appointment lengths “*inadequate for discussing results*”.

#### Assessing DMT eligibility (Supplementary Table 6)

Most GPs (50/80, 62.3%) considered it unlikely that they would begin assessing patient eligibility for DMTs, with free text responses citing a lack of training and it being the remit of specialists (31/56, 55.4%) alongside capacity and funding constraints (15/56, 26.8%).

Respondents did comment that “*stable shared care protocols*” and appropriate guidance (7/56, 12.5%) could favourably influence whether they become involved in assessing patient suitability for medications in the future.

#### Prescribing and monitoring DMTs (Supplementary Table 7 & 8)

Many GPs (27/56, 48.2%) noted that establishing shared care agreements could enable DMT prescribing, with one commenting “*similar to DMARDs [disease-modifying anti-rheumatic drugs] in other specialities would likely need shared care”.* A fifth (11/56, 19.6%) thought this should remain the remit of specialists or that they lacked sufficient training to be able to prescribe. With respect to monitoring DMTs, 17 GPs (32.6%) stated “*we already do this with other medications”* and that monitoring “*is good practice*”.

Shared care agreements were again quoted as facilitators (17/52, 32.6%).

#### Facilitators and barriers (Supplementary Table S &10)

When asked generally about obstacles to the introduction of DMTs (Supplementary Table 9), reservations around timing and capacity were cited in 37/73 (50.7%) narrative responses with one querying “*how could this work possibly fit into primary care in its current form”.* Resourcing and funding were noted as impediments by 32/73 (43.8%) with a GP highlighting the *“limited infrastructure in primary care for infusion or advanced monitoring”.* Others commented on the decision being ICB dependent (13/73, 17.8%) and some raised concerns regarding the rigour of shared care agreements, as evidenced by a response critiquing inconsistencies “*especially when patients move between boroughs or trusts*” and difficulties around “*the lack of adherence to these protocols by secondary care”*.

Comparatively when asked to comment on existing facilitators to the introduction of DMTs (Supplementary Table 10), 19/63 (30.2%) of respondents identified current integrated secondary care and shared care approaches alongside effective and organised practices (10/63, 15.9%) with one participant noting:

*“Integrated Care System (ICS) structures in [region] enable joined up working across primary, community, and secondary care, creating a mechanism to pilot new care models. Electronic care coordination systems such as the Universal Care Plan and shared records through [Egton Medical Information Systems]/Coordinate My Care make it easier to share baseline cognition, comorbidities, and medication data”.*

Most GPs agreed with the survey item (88.8%, 71/80) that more educational instruction is required prior to introduction of DMTs, while 65 explicitly commented on training needs in free text responses (Supplementary Table 11). GPs described needing “*something a bit more extensive than a one hour video lecture, an online module or a quick skim of the NICE guidelines”*, requiring training by experts, protected “*time blocked out to attend”*, and a proper awareness of diagnostics, DMT monitoring and side effects delivered through a blend of online and face to face learning.

## Discussion

### Summary of main findings

Our survey provides a snapshot of primary care perspectives on present and future dementia care. GPs are the initial point of contact for suspected dementia diagnoses in England, though most recorded no direct access to scans despite the inception of community diagnostic centres. Around a third used active monitoring alongside cognitive and blood tests in deciding who to refer to secondary care with presumed dementia. Active monitoring can be a useful and appropriate tool in avoiding referring those unlikely to have dementia onwards, through monitoring for progressive decline, and optimising support for mental and physical health conditions that can engender cognitive symptoms.

Over a quarter of GPs surveyed offered targeted signposting or support for people with MCI, but a quarter were also unaware of specific resources to inform the management of MCI or Functional Cognitive Disorder or said that very little was available. A paucity of research exists regarding how GPs support people who present with cognitive symptoms. As less than half of those with MCI will develop dementia over five years, alternative support to the dementia diagnostic pathway is required (27). Around 10% of people referred to memory services, and 16% of those seen in primary care with cognitive complaints, do not have an objective cognitive deficit (28). People with cognitive symptoms without dementia may benefit from low intensity lifestyle interventions, so there is currently a missed opportunity for secondary dementia prevention (29). Amidst the transfer “from sickness to prevention” GPs will be expected to proactively screen and assist MCI individuals at risk of developing dementia with this highlighted gap in services likely to impede efforts.

GPs provide most post-diagnostic dementia support, with those surveyed collaborating more with voluntary and community organisations. Investment in post diagnostic dementia support within primary care improves patient- and caregiver-related outcomes for people with dementia, when delivered by trained nurses as demonstrated in a German trial (30); similarly, personalised care planning for dementia patients in primary care has shown promise as a sustainable model in England (31). A recent trial of a manualised post-diagnostic dementia support for family carers and people with dementia facilitated by trained and supervised workers without formal clinical training, found that it helped people with dementia achieve personalised care goals cost effectively (32). Only 10 (12.5%) of the practices surveyed had specific provisions for culturally or linguistically underserved groups demonstrating evidence of efforts to personalise care and improve equity of access, yet adopting a more inclusive approach is essential in advance of DMT licensing to prevent widening healthcare inequality (33).

### Strengths and limitations of the study

Strengths of this survey include its iterative development with a range of stakeholders, and the sampling approach we took to mitigate selection biases as far as possible.

There are several limitations. We recruited a broadly representative group of GPs, although the low response rate, probably reflecting the pressure on primary care, is likely to have introduced a response bias towards GPs with a research orientated approach to service planning, an interest in dementia or higher levels of frustration with current systems. Data provided by respondents about their services were not independently verified.

### Comparison with existing literature

Previous research has focussed on attitudes toward early diagnosis, highlighting GP reluctance to formally diagnose dementia within primary care (34,35,36). We instead examined systemic practices that must be altered, reflecting prior research that underscored time, resources and capacity as impediments to adequate primary care dementia support (37). Compared to preceding surveys, we discovered a conditional willingness to adapt, desire to diagnose and minimise waiting times for patients, and for further training to support an enhanced role in diagnostics and support with accompanying renumeration, guidance and protected time. This inclination suggests GPs understand the benefits of their sustained input. There is evidence that higher continuity of GP care for people with dementia is associated with a reduced risk of delirium, incontinence and emergency admissions (38).

### Implications for research and practice

Working in a resource stretched environment, our survey detected apprehensions regarding taking on responsibility for new diagnostic innovations and DMTs due to concerns that despite shared care protocols, greater burden would fall to primary care. Care planning and post diagnostic support will likely remain within this sector aligning with governmental ambitions outlined in the 10 Year Health Plan. However, realising the full potential of a shift from hospital to community requires funding, resource and training needs to be met with the reimagining of relationships between primary, secondary and social care. There are potential advantages to introducing neighbourhood health centres encompassing NHS, social care and community organisations that work cohesively as multidisciplinary teams in local hubs (39). These services could reduce barriers to collaborative working and resource sharing offering a solution to organisational processes that could hamper widespread DMT deployment and may ease primary care burden.

### Funding and Acknowledgments

This research is funded through the NIHR Policy Research Unit in Dementia and Neurodegeneration – Queen Mary University of London, reference NIHR206110. The views expressed are those of the author(s) and not necessarily those of the NIHR or the Department of Health and Social Care.

### Ethics approval

This study was approved by the Health Research Authority and Health and Care Research Wales on 14.5.24 (24/IEC08/0008)(IRAS:338398).

## Data Availability

All data produced in the present study are available upon reasonable request to the author.

