## Supplementary Materials for "A National Survey Exploring Dementia Diagnostics and Care Provisions in Primary Care in England"

### Supplementary Material

**Abbreviations:**
APC: Area Prescribing Committee
CKS: Clinical Knowledge Summaries
CPD: Continuing Professional Development
DMT: Disease Modifying Treatment
GP: General Practitioner
HCA: Health Care Assistant
ICB: Integrated Care Board
MDT: Multi Disciplinary Team Meeting
NHS: National Health Service
NICE: National Institute for Health and Care Excellence
PCN: Primary Care Network
UCP: Universal Care Plan

**Supplementary Figure 1:** Distribution of survey responses from respondents that provided practice level data (Created using Datawrapper [Accessed 22.12.2025] datawrapper.de)

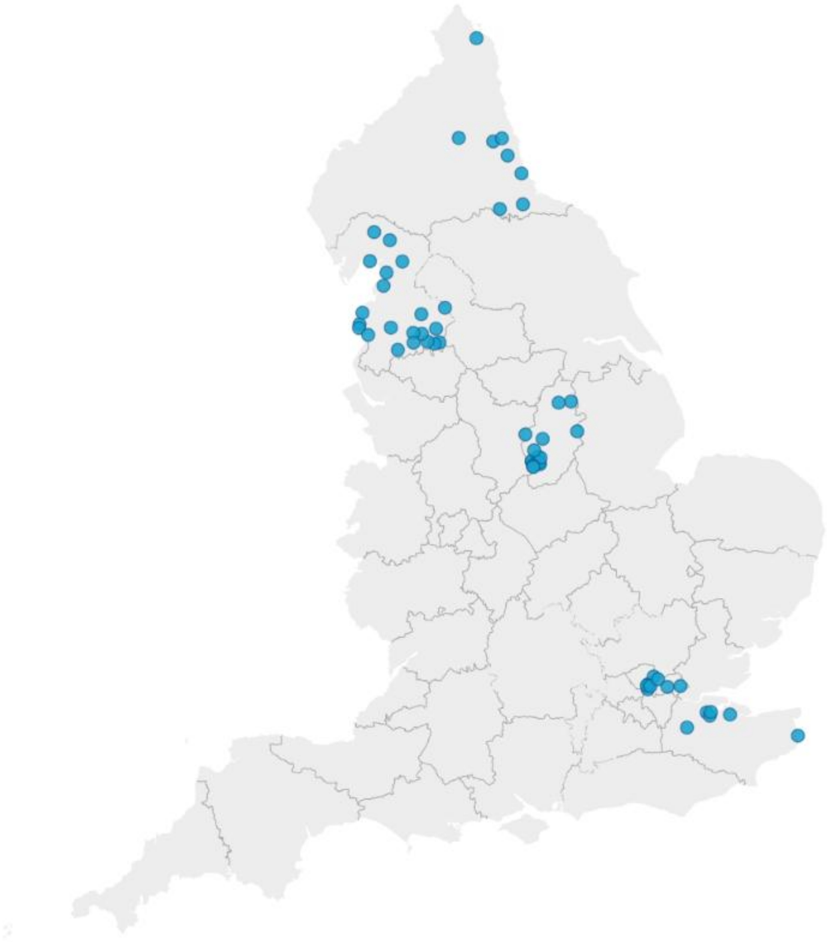

| **Supplementary Table 1:** Content analysis findings of free text responses explaining dementia review processes within GP practices [1] | | |
|---|---|---|
| **Code** | **N = 31 responses** | **Example quotes** |
| **Responsive to individual needs** | 25 (80.6%) | "very patient specific" |
| **Condition-specific** | 8 (25.8%) | "physical health reviews are only if they have other comorbidities like Diabetes"<br>"mental health reviews...case by case... if other mental health diagnoses then yearly or more" |
| **Scheduled reviews** | 8 (25.8%) | "monthly PCN MDT meeting run by a dementia nurse"" |
| **Setting-specific** | 6 (19.4%) | "if in care home then review weekly"<br>"MDT ... every 2 weeks for our care home patients" |
| **Patient or carer initiated** | 3 (9.7%) | "MDTs and Care plans are done if ...initiated by the patient or their carers"<br>"good access so wait for patient initiated appointments" |
| **Practice constraints** | 2 (6.5%) | "looking at these questions is making me feel we don't do enough. But primary care is so squeezed"<br>"we are a very small surgery we don't have many allied professionals" |
| [1] Survey question: "What would you typically do for someone who has just received a diagnosis of dementia? - Other (please specify):" | | |

| **Supplementary Table 2:** Organisations routinely involved in post diagnostic support for patients living with dementia within primary care | |
|---|---|
| **What other organisations are routinely involved in post-diagnostic support for your patients with dementia?** | **N= 80** |
| Clinical memory services (including psychiatry, neurology, geriatric specialities, etc.) | 50 (62.5%) |
| Older adult mental health services | 37 (46.3%) |
| Social care services | 46(57.5%) |
| Voluntary and community sector organisations | 52 (65%) |
| Care Homes | 38(47.5%) |
| None | 7(8.8%) |
| Other: | 13 (16.3%) |
| Social Prescribers | 3(3.8%) |
| Local Integrated Care Team | 1(1.3%) |
| Community/District Nurses | 2(2.5%) |
| Other support Services | 2 (2.5%) |

| **Supplementary Table 3:** Existing guidance, information and resources are available to support GPs in managing subjective and/or functional memory complaints[1] | | |
|---|---|---|
| **Code** | **N= 71 narrative responses** | **Quotes** |
| **CKS/NICE** | 30 (42.3%) | "NICE CKS" |

| | | |
|---|---|---|
| **None/very little/not aware** | 20(28.2%) | "Limited - I'm not aware of any specific resource"<br>"none local that I am aware of" |
| **Specialist team or clinic** | 17(23.9%) | "memory clinic and older age psychiatry team"<br>"Dementia nurse employed by the PCN who has been fantastic" |
| **Online training/general resources** | 15(21.1%) | "in-bedded memory test templates in the clinical system"<br>"generic training resources" |
| **Local/national guidance** | 9(12.7%) | "Nottingham APC guidance" |
| **Charity** | 2(2.8%) | "voluntary services" |
| **Talking therapies** | 2(2.8%) | "NHS talking therapies" |
| [1] "What guidance, information and resources are currently available to support GPs in managing subjective and/or functional memory complaints?" | | |

| **Supplementary Table 4:** Free text responses regarding whether GPs feel their practice may or may not formally diagnose dementia in next 5 years [1] | | |
|---|---|---|
| **Section 1:** Reasons why practices were likely to formally diagnose dementia [2] | | |
| **Code** | **N = 70** | **Example Quote** |
| Systems driven | 15 (22.9%) | "As with many conditions, work is shifting from secondary to primary care"<br>"long delays in memory services mean that diagnosis from Primary Care might be preferential"<br>"Demand for diagnoses outweighing specialist teams ability to do so"<br>"everything is shifting to primary care"<br>"no choice" |
| Personal enthusiasm | 3(4.3%) | "personally as a GP, it would be something I would love to do"<br>"I am hopeful that we can start diagnosing Dementia from the practice itself" |
| Conditional on diagnostic facilities /funding | 3(4.3%) | "access to screening bloods and imaging" |
| Conditional on training/ Support | 2(2.9%) | "with adequate training and supervision" |
| **Section 2:** Reasons why practices were unlikely to formally diagnose dementia[3] | | |
| **Code** | **N= 70** | **Example Quote** |
| Lack of confidence/ expertise/ training | 32(45.7%) | "GPs not fully trained in diagnosis"<br>"it requires expertise from an old-age psychiatrist to complete this diagnose"<br>"I feel needs specialist assessment" |

| | | |
|---|---|---|
| Capacity/funding | 21(30%) | "This is a complex diagnosis that can be mimicked by other mental and physical health problems. It is a diagnosis made after a number of appointments and use of longer memory assessments than we are able to do in primary care"<br>"funding is the key"<br>"present to capacity at GPs to see lot of acutely unwell patients. Unless more GP surgeries are opened no extra work can be taken" |
| Lack of diagnostic facilities/tests | 9(12.9%) | "don't have access to imaging" |
| Systems-reasons | 7(10%) | "depends on services and ICB decisions"<br>"need ICB permission to complete assessment for formal diagnosis"<br>"depends on services and ICB decisions - current waiting list for memory clinic in Nottingham is ~7-8 months" |
| [1]Survey Question: **"**In the next 5 years do you expect your practice to consider formally diagnosing dementia? Please can you briefly explain why."<br>[2]22/70 respondents who left free text responses mentioned facilitators<br>[3] 49/70 respondents mentioned barriers | | |

| **Supplementary Table 5:** Free text responses exploring why blood biomarkers may or may not be offered by GPs within primary care in the next five years [1] | | |
|---|---|---|
| **Section 1:** Reasons why practices are likely to consider taking blood samples for dementia biomarker testing [2] | | |
| **Code** | **N=57 responses** | **Example Quotes** |
| **Could become part of routine screenings/ willing to adopt** | 25(43.9%) | "very much welcomed as it will give us a solid evidence to diagnose and treat within primary care"<br>"relatively easy change to make to our practice" |
| **With funding/ access** | 20(35.1%) | "All depends on whether the lab makes this available"<br>"Depending if NHS funds it"<br>"think this depends on local availability" |
| **With support/ training/shared care** | 11(19.3%) | "with the right guidance this is something we can initiate"<br>"under a shared care agreement" |
| **Already routinely take bloods as part of dementia screen** | 6(10.5%) | " have a phlebotomy clinic with 2 HCAs ... routinely take bloods for dementia referrals and as part of the MMSE" |
| **If recommended** | 2(3.5%) | "If recommended in guidance" |

| **Section 2:** Reasons why practices are unlikely to consider taking blood samples for dementia biomarker testing [3] | | |
|---|---|---|
| **Code** | **N=57 responses** | **Example Quotes** |
| **Capacity/funding** | 8(14.0%) | "Cost is too high"<br>"Where would the funding come from for this" |
| **Unavailable/ accessibility barriers** | 6(10.5%) | "not available"<br>" I highly doubt they'd let lowly GPs have access" |
| **Remit of specialists** | 5(8.8%) | "Why should this burden fall on GPs when there are secondary care services commissioned to diagnose dementia"<br>"Understanding when to test/interpreting results of novel biomarkers is likely not within our remit, this should come through secondary care" |
| **Training concerns** | 5(8.8%) | "Not trained in interpretation of these" |
| **Lack of knowledge/awareness of tests** | 3(5.3%) | "Unfamiliar with these tests" |
| [1] "Survey Question: In the next 5 years, do you expect your practice to consider taking blood samples for dementia biomarker testing? Please can you briefly explain why."<br>[2] 38/57 participants that left free text responses mentioned facilitators<br>[3] 20/ 57 respondents mentioned barriers | | |

| **Supplementary Table 6:** Free text responses regarding whether GPs may or may not assess their patients' eligibility for DMTs in next 5 years [1] | | |
|---|---|---|
| **Section 1:** Reasons why practices were likely to assess patient eligibility for DMTs [2] | | |
| **Code** | **N = 56** | **Example Quote** |
| Conditional on diagnostic shared care or support | 7 (12.5%) | "would want final decision to be shared- maybe in a MDT" |
| Within GP remit/ shift from secondary to primary care | 5(8.9%) | "As we would assess suitability for all medications"<br>"We see/know the patients and families/carers very frequently, we understand what is important/matters for the specific patient and family and how the condition is progressing"<br>"I can easily foresee the ICB/NHSE off-loading this to GPs to" |
| If commissioned | 3(5.4%) | "if prescribing committee allows if in primary care" |

| | | |
|---|---|---|
| Willingness to | 2(3.6%) | “Happy to be part of the assessment process” |
| With funding/ resources | 2(3.6%) | “Depends on remuneration”<br>“will depend on local funding” |
| **Section 2:** Reasons why practices were unlikely to assess eligibility for DMTs[3] | | |
| **Code** | **N= 56** | **Example Quote** |
| Remit of specialist or secondary care/ lack of training or expertise | 31(55.4%) | “Pretty busy in primary care! Specific drugs with limited experience, or drugs with higher than average risks are still generally initiated in secondary care”<br>“That is the role of MHSOP” |
| Capacity/funding issues | 15(26.8%) | “Will be too pricey initially for NICE”<br>“Would need significant investment in resourcing ... If GPs are going to be doing all this what are the funded memory services going to be doing?” |
| System dependent | 3(5.6%) | “this isn’t a decision we make” |
| [1]Survey Question**: “**In the next 5 years do you expect your practice to consider assessing patient’s eligibility for DMTs for dementia? Please can you briefly explain why.<br>[2]15/ 56 respondents who left free text responses mentioned facilitators<br>[3] 41/56 respondents mentioned barriers | | |

| | | |
|---|---|---|
| **Supplementary Table 7:** Free text responses regarding whether GPs may or may not prescribe dementia DMTs to their patients in next five years[1] | | |
| **Section 1:** Reasons why practices are likely to prescribe dementia DMTs [2] | | |
| **Code** | **N = 56** | **Example quote** |
| With a shared care agreement | 27(48.2%) | “once initiated under shared care agreement”<br>“we already do take over prescribing of these meds once the patients are stable on them in secondary care” |
| If commissioned | 9(16.1%) | “the local prescribing committee believe it is safe to do so in primary care”<br>“depends on what is commissioned” |
| With training or support | 6(10.7%) | “With the correct training and support”<br>“with appropriate support and guidance this could be possible” |
| With requisite funding, renumeration or resources | 6(10.7%) | “contractual remuneration, regulations needed to be met”<br>“needs proper funding to primary care as you keep throwing secondary care work on us”<br>“with ... resourcing it is likely that in straight forward cases this would be appropriate work in GP” |

| | | |
|---|---|---|
| Shift from secondary to primary care | 1(1.8%) | "Cheaper to get a GP to do it" |
| **Section 2:** Reasons why practices are unlikely to prescribe dementia DMTs[3] | | |
| **Code** | **N= 56** | **Example quote** |
| Remit of specialist or secondary care/ lack of training or expertise | 11(19.6%) | "lack of skills"<br>"not within remit of general practice"<br>"local gps are not trained to prescribe disease modifying drugs" |
| Capacity/funding issues | 6(10.7%) | "Currently we are not accepting unresourced shared care agreements"<br>"no funding"<br>"not paid for this work" |
| No current guidance | 2(3.6%) | "Not received any information about this"<br>"No current guidelines for this in primary care" |

[1]Survey question**: "**In the next 5 years do you expect your practice to consider prescribing disease modifying treatments for dementia? Please can you briefly explain why."
[2]15/ 56 respondents who left free text responses mentioned facilitators
[3] 41/56 respondents mentioned barriers

**Supplementary Table 8:** Free text responses regarding whether GPs may or may not monitor the progress and side effects of dementia DMTs in the next five years [1]

| | | |
|---|---|---|
| **Section 1:** Reasons why practices are likely to monitor the effects of dementia DMTs [2] | | |
| **Code** | **N = 52** | **Example quote** |
| With a shared care agreement | 17(32.6%) | "once initiated under shared care agreement"<br>"we already do take over prescribing of these meds once the patients are stable on them in secondary care"<br>"I would expect a patient to be stable on a drug before the prescribing is transferred to primary care. We would then monitor at advised intervals for side effects and expect support from secondary care where there are any concerns" |
| Already responsible for medication monitoring/ GPs role | 17(32.6%) | "we already do this!"<br>"in reality [memory clinics] will be inaccessible to patients who will ask GP to review if they have side effect"<br>"we do this with multiple other drugs and conditions" |
| With training or support | 7(13.5%) | "With the correct training and support"<br>"with appropriate support and guidance this could be possible"<br>"with correct guidance and training we can" |

| | | |
|---|---|---|
| Depending on resources/ capacity | 3(5.8%) | "In the future if there is a significantly bolted Primary and community care workforce that included clinical prescribers and colleagues who were competent are dealing with specific medication side effects as they are" |
| Shift from secondary to primary care | 1(1.9%) | "Lots of other specialities already off-load this care to GPs" |
| **Section 2:** Reasons why practices are unlikely to monitor the effects of dementia DMTs[3] | | |
| **Code** | **N= 52** | **Example quote** |
| Remit of specialist or secondary care/ lack expertise | 7(13.5%) | "This is a specialist area. - not within the remit of general practice"<br>"should be done by secondary care" |
| Capacity/funding issues | 7(13.5%) | "No time in primary care"<br>"If it involves long memory assessments and input then with the current pressure on primary care then this would be difficult to achieve" |
| Need training | 1(1.9%) | "our current knowledge on this is low - what do we do to manage side effects, doses, who do we get advice and support from?" |
| [1]Survey question: **"**In the next 5 years do you expect your practice to consider monitoring progress and side effects of disease modifying treatments for dementia? Please can you briefly explain why."<br>[2]35/ 52 respondents who left free text responses mentioned facilitators<br>[3] 13/52 respondents mentioned barriers | | |

| **Supplementary Table 9:** Barriers to GP practices introducing dementia DMTs [1] | | |
|---|---|---|
| **Code** | **N= 73 responses** | **Example quote** |
| **Time/capacity issues** | 37 (50.7%) | "How could this work possibly fit into primary care in its current form" |
| **Training** | 34(46.6%) | "knowledge"<br>"No barriers. We clinicians need training" |
| **Resourcing/funding** | 32 (43.8%) | "Doing this work will inevitably mean our resources to do other areas of work are further stretched"<br>"Limited infrastructure in primary care for infusion or advanced monitoring" |

| | | |
|---|---|---|
| **ICB/Commissioner approval** | 13(17.8%) | "Need to be approved by ICB - unlikely as they still have not approved weight loss injections despite government policy for all regions to do so"<br>"Neighbourhood health's apparent focus on the frail will possibly steer away from facilitating diagnosis of the younger pts who may benefit most" |
| **Poor previous shared care agreements/remit of secondary care** | 8(11%) | "Established shared care protocols in other disease areas (!) The lack of adherence to these protocols by secondary care and the assumption that GP practices should pick up the slack to benefit patients even when this is dealing with things quite clearly outside of our expertise."<br>"shared care across multiple providers still inconsistent, especially when patients move between boroughs or trusts"<br>"lack of appetite for taking on work from secondary care" |
| **Equal access concerns** | 3(4.1%) | "Equity and access concerns...accessing new treatments"<br>"Distance by foot or public transport from the local hospital... lack of safe local spaces for these patients to leave their homes"<br>"difficult for pts to access primary care in the first place" |

| | | |
|---|---|---|
| **Low demand** | 2(2.7%) | "We have very few patients with dementia (skewed demographic)" "small patient numbers" |
| [1] "Please describe any barriers to the introduction of new disease modifying treatments for dementia to your practice" | | |

| **Supplementary Table 10:** Existing facilitators of DMT adoption within primary care[1] | | |
|---|---|---|
| **Code** | **N =63 responses** | **Quotes** |
| **None/not aware** | 23(36.5%) | "none at present" |
| **Integrated secondary care/ shared care** | 19(30.2%%) | "close working relations with our local memory clinic" "Established multi-disciplinary links with community mental health teams, memory services" |
| **Good clinical staff** | 12(19.0%) | "excellent clinical pharmacist" |
| **Effective practice/organisation readiness** | 10(15.9%) | "Electronic care coordination systems such as the Universal Care Plan and shared records through EMIS/Coordinate My Care make it easier" "Our practice maintains an up-to-date dementia register with active recall and monitoring processes, allowing efficient identification of eligible patients for disease-modifying treatments" |
| **Training** | 7(11.1%) | "Training. Allocating of dementia lead" |
| **Willingness to adopt** | 4(6.3%) | "would myself be willing to embark and implement such a treatment in my practice" |
| **Funding** | 3(4.8%) | "funding" |
| [1] "Please describe any existing facilitators to the introduction of new disease modifying treatments for dementia to your practice" | | |

| **Supplementary Table 11:** Free text responses outlining training needed before integration of dementia DMTs[1] | | |
|---|---|---|
| **Code** | **Number = 65 narrative responses** | **Example quotes** |
| **Training on DMTs, monitoring & side effects** | 29(44.6%) | "red flags to look out for" |
| **Unspecified training** | 22(33.8%) | "all"<br>"guidance adequate to primary care settings" |
| **Diagnostics** | 21(32.3%) | " identifying different dementias and differentials from other conditions which may mimic dementia, when to image, when to check blood tests"<br>" Understanding of results of bio marker tests and imaging" |
| **Online learning/webinars** | 9(13.8%) | "online accessible at any time with CPD" |
| **Capacity concerns/should remain in specialist secondary care** | 7(10.7%) | "Concerns about over diagnosis, capacity"<br>"I argue this should primarily be the job of secondary care." |
| **In person** | 7(10.7%) | "actual sit in clinics or workshop" |
| **Training by experts/Access to specialist support** | 6(9.2%) | " Something a bit more extensive than a one hour video lecture, an online module or a quick skim of the NICE guidelines...contact details for clinical advice"<br>"access to specialist advice when we get stuck"<br>"Clear contact details for helpline/email address for clinical advice/troubleshooting" |
| **Funding concerns/ renumeration needed** | 5(6.7%) | "remuneration for staff time to attend the training, backfill locum costs for the clinicians to be relieved of their current duties" |

| | | |
|---|---|---|
| **Protected time** | 3(4.6%) | “time blocked out to attend” |
| [1] **“**What training do you feel would be most helpful?” | | |

| Topic | Questions |
| --- | --- |
| 1. Pre-diagnostic dementia care | What do you do if you suspect that someone has dementia?<br>If you use cognitive screening/psychometric tests. Which type of screening tool do you typically use?<br>Does your practice have access to dementia specific blood biomarker testing (e.g. tau or amyloid)?<br>If you request neuroimaging, what type of neuroimaging do you use?<br>Does your practice have direct access to neuroimaging?<br>If you refer onwards who do you refer to?<br>Approximately how many people have you referred for a dementia assessment in the last year? |
| 2. Post diagnostic care | What would you typically do for someone who has just received a diagnosis of dementia?<br>For patients with dementia how often does your practice conduct:<br>Care plan reviews<br>Mental health reviews<br>Medication reviews<br>Physical health reviews<br>Multi-disciplinary team meetings?<br>What other organisations are routinely involved in post diagnostic support for your patients with dementia?<br>Does your practice offer or signpost any specific support for people with:<br>Dementia from minority communities?<br>Mild Cognitive Impairment?<br>Parkinson’s disease and cognitive impairment? |
| 3. Service outcomes and future planning | In the next 5 years, do you expect your practice to consider:<br>Formally diagnosing dementia? (Please can you briefly explain why)<br>Taking blood samples for dementia biomarker testing? (Please can you briefly explain why)<br>Assessing patient’s eligibility for disease modifying treatments for dementia? (Please can you briefly explain why)<br>Prescribing disease modifying treatments for dementia? (Please can you briefly explain why)<br>Monitoring progress and side effects of disease modifying treatments for dementia? (Please can you briefly explain why)<br>Do you feel that you need any further training to prepare for these challenges?<br>What training do you feel would be most helpful?<br>Please describe any existing facilitators to the introduction of new disease modifying treatments for dementia to your practice.<br>Please describe any barriers to the introduction of new disease modifying treatments for dementia to your practice. |

| | |
|---|---|
| | What guidance, information and resources are currently available to support GPs in managing subjective and/or functional memory complaints? |
| 4. Participant demographics and practice characteristics | How many years have you worked as a GP?<br>What is your age?<br>Which gender do you most identify with?<br>What is your ethnic group?<br>How big is your practice by patient list size?<br>How would you describe your practice geographically?<br>How big is your practice in terms of number of GPs?<br>In which Integrated Care Board (ICB) is your practice located?<br>In which Primary Care Network (PCN) is your practice located?<br>What is the name of your practice? |
| **Box 1:** Overview of survey questions | |
